# Life Expectancy and Voting Patterns in the 2020 U.S. Presidential Election

**DOI:** 10.1101/2020.10.26.20220111

**Authors:** Lesley H. Curtis, Molly N. Hoffman, Robert M. Califf, Bradley G. Hammill

## Abstract

**Introduction:** In the 2016 U.S. Presidential election, voters in communities with recent stagnation or decline in life expectancy were more likely to vote for the Republican candidate than in prior Presidential elections. We aimed to assess the association between change in life expectancy and voting patterns in the 2020 Presidential election.

**Methods:** With data on county-level life expectancy from the Institute for Health Metrics and Evaluation and voting data from GitHub, we used weighted multivariable linear regression to estimate the association between the change in life expectancy from 1980 to 2014 and the proportion of votes for the Republican candidate in the 2020 Presidential election.

**Results:** Among 3,110 U.S counties and Washington, D.C., change in life expectancy at the county level was negatively associated with Republican share of the vote in the 2020 Presidential election (parameter estimate −7.2, 95% confidence interval, −7.8 to −6.6). With the inclusion of state, sociodemographic, and economic variables in the model, the association was attenuated (parameter estimate −0.8; 95% CI, −1.5 to −0.2).

**Conclusion:** Counties with a less positive trajectory in life expectancy were more likely to vote for Republican candidates in the 2020 U.S. Presidential election, but the association was mediated by demographic, social and economic factors.

## Introduction

Although life expectancy in the U.S. increased by nearly 10 years over a nearly 60-year period from 1959 to 2016, the rate of increase slowed over time and life expectancy has declined after 2014.^1^ Large disparities in life expectancy among counties in the U.S. have increased as well. In 2014, a 20-year gap existed between counties with the lowest and highest life expectancy. A gap of more than 10 years in life expectancy existed between counties in the 1st and 99th percentile, an increase of 2.4 years from 1980.^2^

Recent reports have described the association between changes in life expectancy and voting behavior, noting a shift toward the Republican presidential candidate in counties where life expectancy has declined or increased the least.^3-5^ One analysis found a 9.1 percentage point increase in the Republican vote share from 2008 to 2016 in counties in which life expectancy stagnated or declined from 1980 to 2014.^4^ Voters in those counties were more likely to vote for Republican candidates in 2016, irrespective of prior voting patterns.

Following the 2020 Presidential election, we examined the election results to determine whether the pattern observed in 2016 persisted. In this brief report, we examine the association between change in county-level life expectancy from 1980 to 2014 and the proportion of votes cast for the Republican party in the 2020 U.S. Presidential election.

## Methods

We obtained county-level data on life expectancy from the Institute for Health Metrics and Evaluation^6^ and calculated the absolute difference in life expectancy from 1980 to 2014. We obtained county-level voting results from the 2020 Presidential election from GitHub^7^, which were scraped from results published by Fox News, Politico, and the New York Times, and calculated the Republican candidate’s share of votes as the proportion of total votes cast for the Democratic and Republican candidates in each county and Washington, D.C.. Across 3,109 counties with any votes not attributed to the Democratic or Republican parties, the median percentage of votes for the third-party candidate was 2%, so we excluded votes for third-party and other candidates from the denominator. We also excluded one county due to missing data and counties in Alaska and Kalawao County, Hawaii due to a discrepancy in county classifications between the data sources, so 3,110 counties and Washington, D.C. were included in the final analysis.

We used multivariable linear regression weighted by the number of two-party votes to assess the association between the absolute change in life expectancy from 1980 to 2014 and the proportion of votes for the Republican candidate in the 2020 Presidential election. The multivariable model included adjustments for state, urban-rural classification using the 2013 NCHS Urban-Rural Classification Scheme for Counties^8^, and the following sociodemographic and economic characteristics from the 2015-2019 5-year American Community Survey: percent college educated, Gini coefficient, unemployment rate, median house value, poverty rate, percent Black, and percent Hispanic.^9^ Median home value was transformed to the logarithmic scale and percent Black and Hispanic were transformed to their square root to address skewed data. We used heteroskedasticity-consistent standard errors to calculate 95% confidence intervals. We generated maps of the national distribution of voting patterns and life expectancy and scatter plots displaying the relationship between change in life expectancy and voting patterns. This study was determined exempt by the Duke Health Institutional Review Board.

## Results

From 1980 to 2014, the change in county-level life expectancy ranged from −2.2 years to 10.8 years with a median change of 3.9 years (inner quartile range [IQR] 3.0 to 4.8). (Figure 1) The county-level proportion of the two-party vote for the Republican candidate in the 2020 Presidential election ranged from six percent to 97% with a median of 70%. (Figure 2). After weighting for the total number of two-party votes, the weighted median percent of the two-party vote for the Republican candidate was reduced to 46% (IQR 36 to 60%).

**Figure 1:**
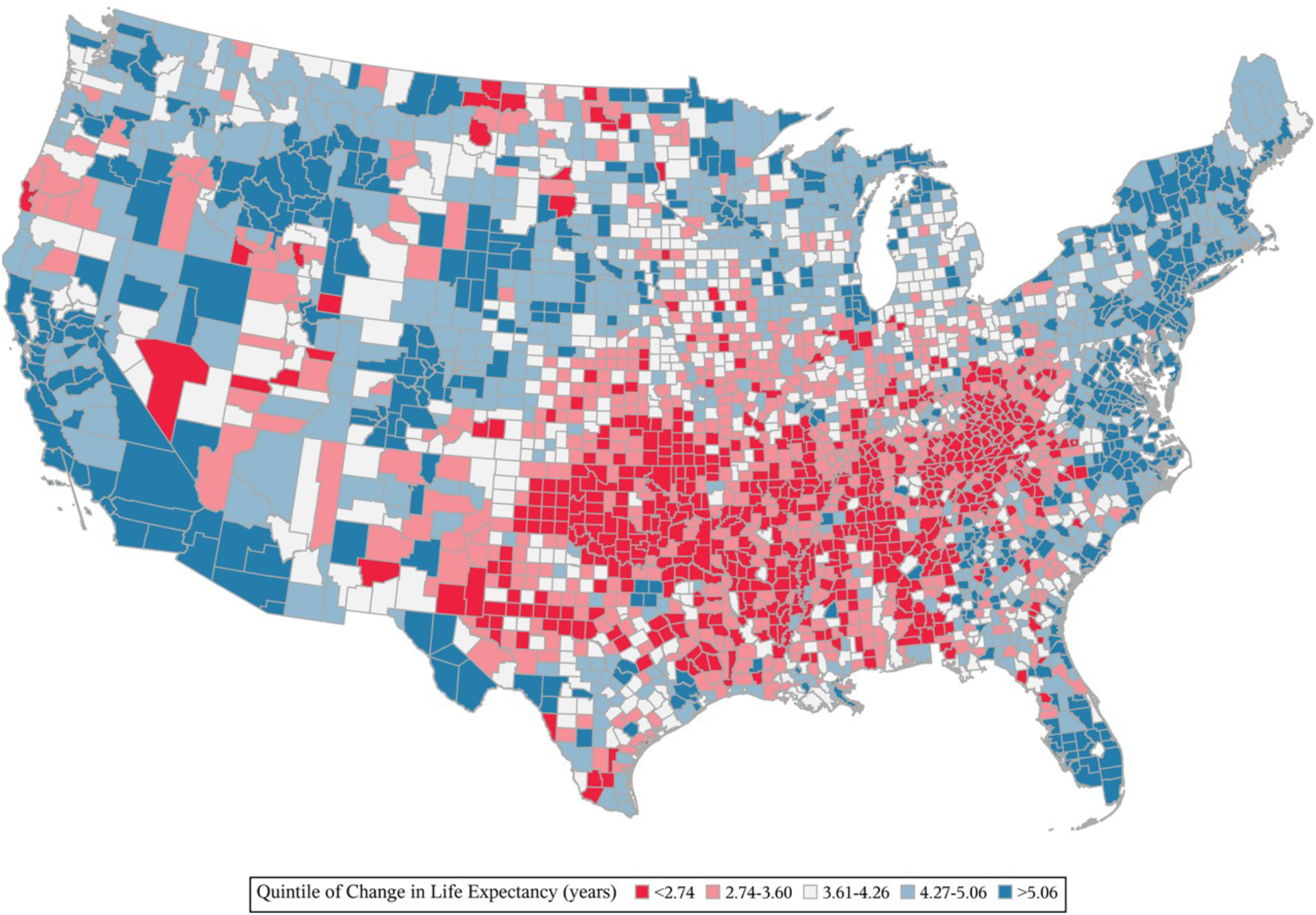
Change in life expectancy in U.S. counties from 1980 to 2014

**Figure 2:**
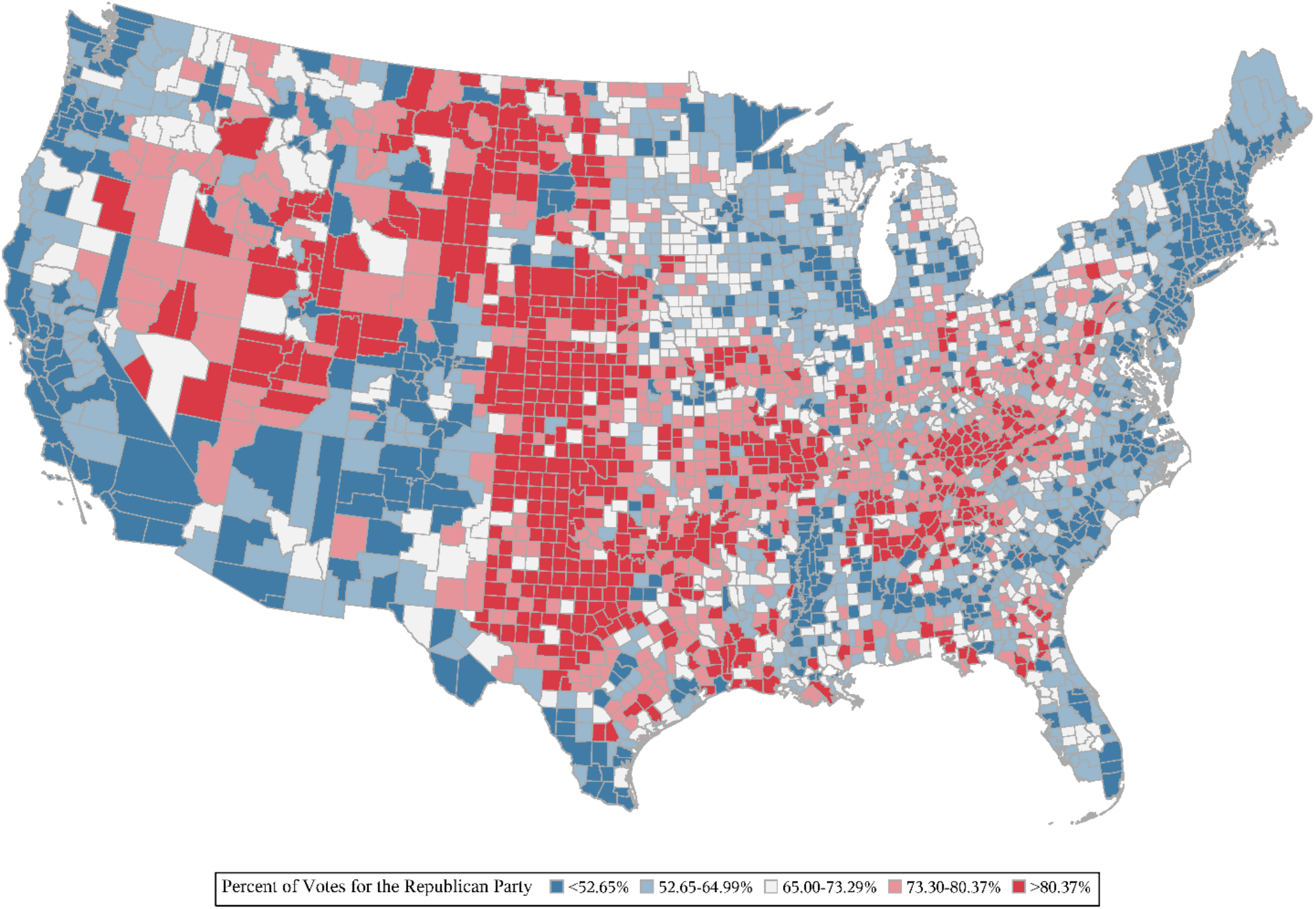
Republican Candidate’s Share of the Vote in the 2020 U.S. Presidential election.

Change in life expectancy at the county level was negatively associated with Republican share of the county-level vote in the 2020 Presidential election. For each year of increase in life expectancy, the Republican vote share at the county level decreased by 7.2 percentage points (95% CI, −7.8 to −6.6). (Figure 3) With the inclusion of state, sociodemographic, and economic variables in the model, the association was attenuated (parameter estimate −0.8; 95% CI, −1.5 to −0.2) (Figure 4 and Table 1).

**Table 1:**
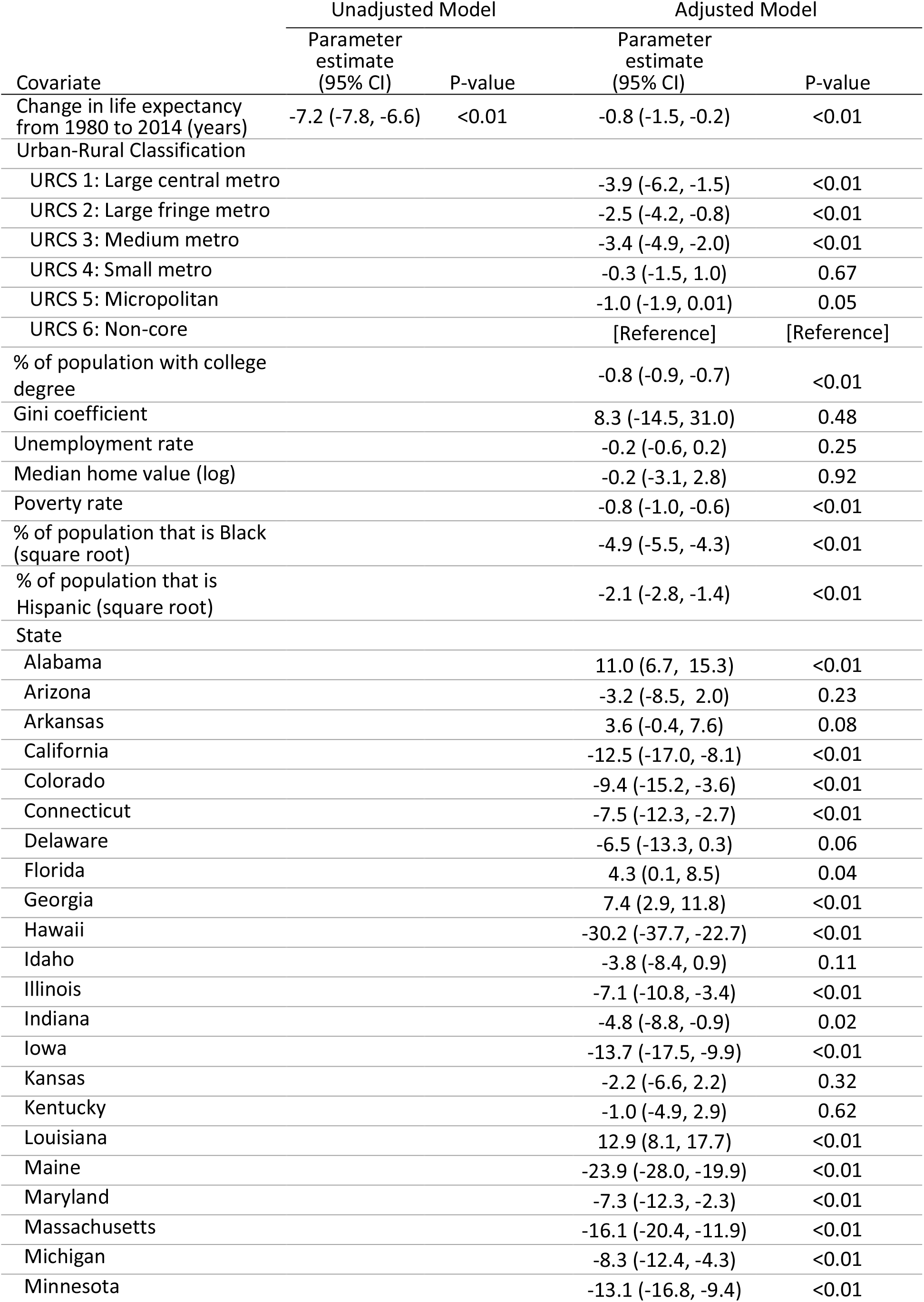
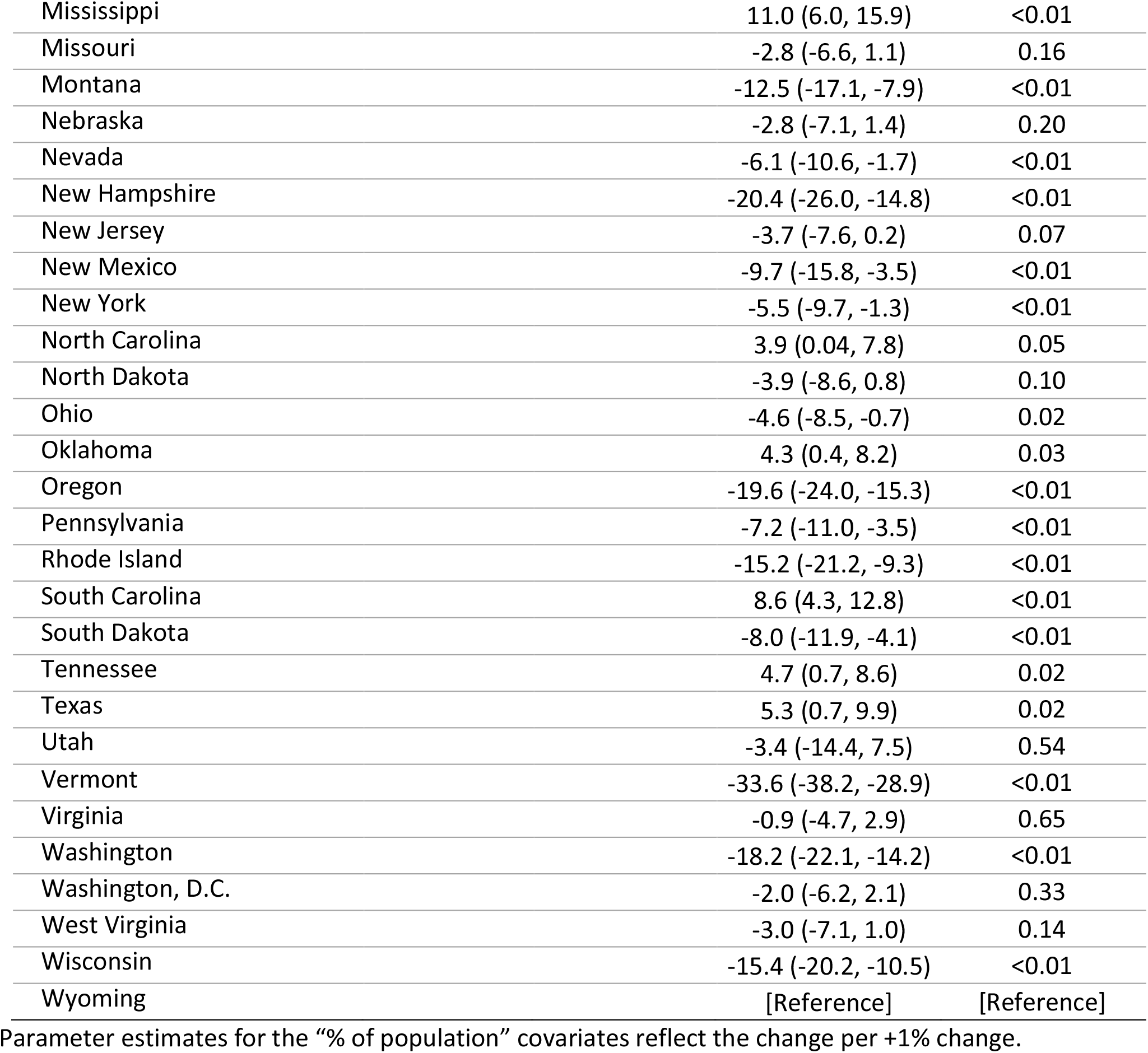
Association Between Change in County-Level Life Expectancy from 1980 to 2014 and Republican Share of the County-level Vote in the 2020 Presidential Election

**Figure 3.**
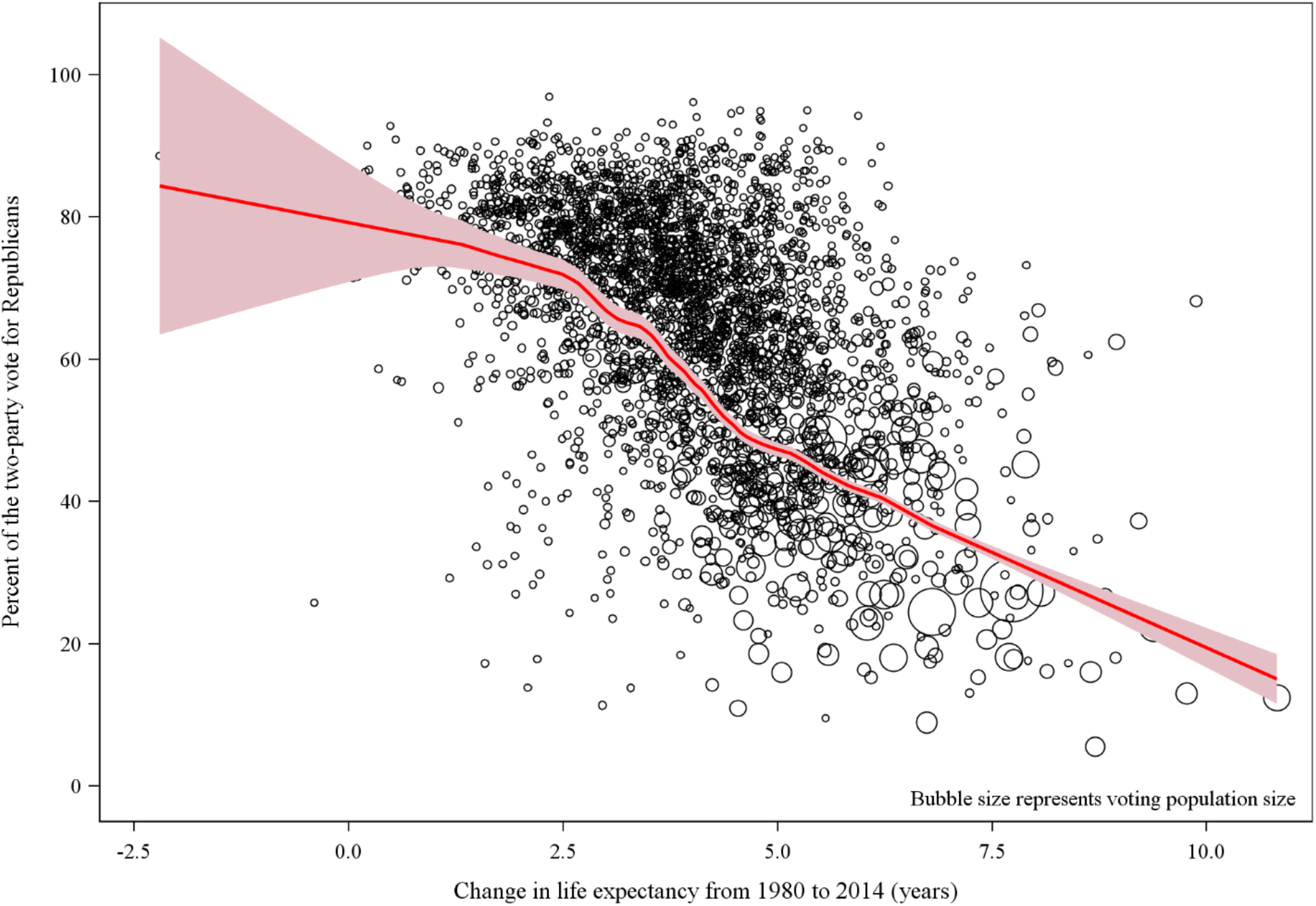
Relationship Between Change in County-Level Life Expectancy and Republican Share of the County-level Vote in the 2020 Presidential Election

**Figure 4.**
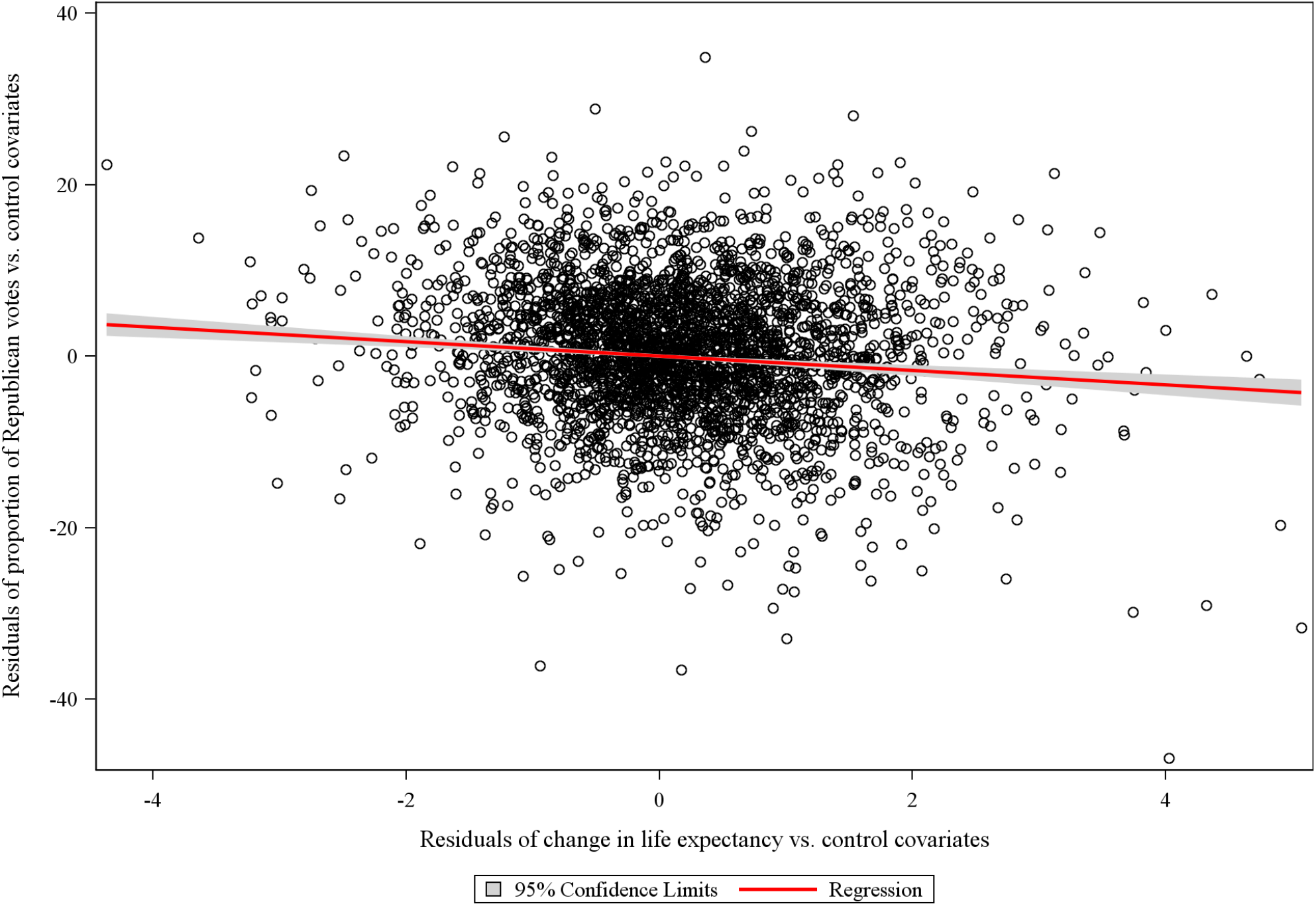
Residuals of Change in Life Expectancy vs. Control Covariates and Residuals of Proportion of Republican Votes vs. Control Covariates

## Discussion

Compared to prior elections, the 2016 U.S. Presidential election witnessed a shift of votes toward the Republican candidate in communities that have experienced recent stagnation or decline in life expectancy.^4,5^ A recent analysis examined the association between community health and the shift from 2016 to 2018 in Republican share of the vote in U.S. House of Representatives elections.^10^ Although there was a general shift toward the Democratic candidate in the 2020 election, we found that voting in less healthy communities continued to favor the Republican candidate. When we adjusted for demographic, social and economic factors, the difference was attenuated, suggesting that these factors mediated the observed association between change in life expectancy and voting status.

Compared with other high-income countries, the U.S. has the lowest life expectancy with a trajectory that is diverging from other high-income countries.^11^ In addition, substantial disparities in life expectancy as a function of geographic location are well documented within the U.S. Figures 1 and 2 offer graphic visual representation of the congruence of life expectancy and voting patterns. While this epiphenomenon is interesting, our analysis suggests that community voting patterns are likely to represent manifestations of differences in demographics, economic well-being, and educational status. Regardless of which political party is in power, effective policies must be implemented to improve health outcomes in counties and regions where life expectancy has stagnated.

This study is limited by its ecological nature and the inability to associate individual voting behaviors with health outcomes. Additionally, the study is limited by available data sources and would benefit from more detailed social and economic data. Updated estimates of life expectancy will be critical to guide policies to improve the well-being of negatively affected counties, but these data are not yet available.

In conclusion, counties with a less positive trajectory in life expectancy were more likely to vote for the Republican candidate in the 2020 U.S. Presidential election. The association was mediated by demographic, social and economic factors that should drive health policy priorities over the coming years.

## Data Availability

All data used in the manuscript are publicly available. Links are provided below and referenced in the manuscript.

https://github.com/tonmcg/US_County_Level_Election_Results_08-20

https://www.cdc.gov/nchs/data_access/urban_rural.htm#Data_Files_and_Documentation

https://www.census.gov/programs-surveys/acs/data.html

http://ghdx.healthdata.org/record/ihme-data/united-states-life-expectancy-and-age-specific-mortality-risk-county-1980-2014

